# Variation in Mycobacterium tuberculosis genotype and molecular phenotype influence clinical phenotype of Pulmonary tuberculosis and Tuberculous Meningitis infection in host

**DOI:** 10.1101/2020.11.05.20225789

**Authors:** Aliabbas A. Husain, Hitesh Tikariha, Amit R. Nayak, Umesh D. Gupta, Shraddha S. Bhullar, Nitin C. Chandak, Tanya M. Monaghan, Hatim F. Daginawla, Lokendra R. Singh, Hemant J. Purohit, Rajpal S. Kashyap

**Author notes:** Corresponding author Rajpal S. Kashyap, **E-mail:**.

## Abstract

In the present study, we investigated tissue specific colonization and virulence characteristics of two different Mycobacterium tuberculosis (MTB) clinical isolates derived from patients with Pulmonary TB (PTB) and Tuberculous Meningitis (TBM). We retrospectively studied a total of 1458 patients diagnosed with TB between 2003 and 2013. Of these, archived sputum and CSF samples were available for 323 TBM and 157 PTB patients. We selected a total of 10 sputum and CSF isolates from each group for further molecular characterization. Methodologies employed included, Gas chromatographic analyses of Fatty Acid Methyl Esters (GC-FAME) followed by MTB genotyping to assess strain diversity. We further assessed the molecular phenotype of each strain through in-vitro cytokine assays & murine MTB model. Our comparative genomics data illustrated two diverse genotypes belonging to H_37_R_V_ (PTB) and CCDC5079 linkage (TBM), highlighting major variation in membrane protein composition and enzymes that make different mycolic acids. The differential cytokine response by both the strains & GC-FAME analysis further corroborated this variation in membrane composition. This was in agreement with KasIII enzyme, LppA and desaturase related variation in protein. Both MTB strains in mice showed diverse pathogenesis with CCDC5079 infected mice exhibiting higher dissemination to brain compared to the H_37_R_V_ strain which developed progressive pulmonary disease. These observations suggest that variation in the MTB membrane composition could play an important role in differential colonization of these strains. The study warrants further investigation of membrane proteins with respect to blood brain barrier invasion and pathogenesis.

## 1. Introduction

Tuberculosis (TB) is a major public health problem in India, accounting for over a quarter of the global TB and multidrug-resistnt TB (MDR-TB) cases [1]. Although pulmonary TB (PTB) is the primary manifestation of *Mycobacterium*. *tuberculosis* (MTB) infection, there has been an exponential rise in the number of extra-pulmonary TB (EPTB) cases in India over the last two decades. Among EPTB, Tuberculous Meningitis (TBM) is the most severe, accounting for 70-80% all neurological TB cases and is often associated with a high rate of neurological sequelae [2].

As reported earlier by various groups, TBM is often preceded by a respiratory infection followed by early haematogenous dissemination to central nervous system (CNS) [3-4] with one report indicating that pulmonary manifestations precede onset of neurological symptoms in 75% of patients [5]. In a our previous retrospective analysis of CNS infections in our hospital between 2000-2011 we diagnosed 2000 cases of TBM. Interestingly, 55% of these cases presented with clinical manifestation of TBM without miliary TB (all patients had no active pulmonary tuberculosis when the meningitis was diagnosed) [Kashyap et al, un published data]. In these TBM patients, there was the distinct absence of MTB mediated immune response in blood. Furthermore, radiological investigations of lungs were not suggestive of TB, possibly indicating lack of pulmonary foci in TBM infection which is in agreement with earlier reports which suggest 25-30% of TBM cases do not have pulmonary involvement. [5]

It remains unclear how MTB directly affects the meninges of the brain without any haematogenous involvement. Evidence from previous animal studies indicates that certain MTB strains evoke a different immune-pathologic host response due to variation in virulence factors, and therefore these strains may exhibit a specific ability to invade the CNS blood brain bar rier (BBB) and cause disseminated disease [6-7]. Although various MTB strains have been characterized and reported [8-9], limited studies exisit on variation MTB genotype and their relevance with respect to disease outcome in host.

In the present work, we report the comprehensive molecular characterization of two MTB isolates recovered from PTB and TBM cases (hereafter referred as S3 and C3) and their strain virulence and pathogenesis using in-vitro cell line and murine model of MTB infection. The major aim of was to study whether variation in MTB genotype or phenotype is responsible for differential site specific colonization in host leading to different forms of TB upon infection.

## 2. Material and Methods

### 2.1 Ethics statement

All studies on human subject including collection of sputum and CSF samples from PTB and TBM cases were approved by Ethical Committee of Dr. G. M. Taori Central India Institute of Medical Sciences (CIIMS), Nagpur. Informed consents were obtained from all particpants after oral explanation of the study to them. All protocols for animal experiments, including infection, dosing were approved by Ethical committee of CIIMS and National JALMA Institute of leprosy and other mycobacterial diseases, Agra (JALMA), Uttar Pradesh, India.

### 2.2 Study setting and participant recruitment

An clinical observational and molecular study was conducted at CIIMS, Nagpur, a frontline tertiary care neurology hospital in Central India. The hospital serves approximately 60000 patients per year, 70% of which are neurological cases. A total of 1458 patients with TB admitted to CIIMS from 2003 to 2013 were included in the study using predefined clinical inclusion criteria as per standards of TB care in India [10]. For each medical case file, the patient’s history, physical findings, radiographical information and reports of laboratory investigations were assessed to obtain the necessary information relating to diagnosis of TB. For each patient, demographic information (age, gender), lifestyle factors (smoking habits and alcohol use) and clinical characteristics were recorded. Clinical characteristics included, co-morbid conditions (diabetes mellitus, hypertension, malignancies, other infections encountred in the past one year), previous treatment for TB, and history of known contact with an active TB case. Radiological investigations included chest radiographs and computerized tomography (CT) of PTB and TBM cases.

Of the 1458 cases eligible for inclusion, 632 TBM cases died, emigrated or were lost to follow-up within the first year of diagnosis. 68 cases did not give consent for the study. 81 cases were excluded due to lack of sample Cerebrospinal fluid (CSF) or sputum for analysis. 677 cases of TB were included in the final analysis. Among these, 144 cases were diagnosed with bacterial meningitis and thus excluded. Of the remaining 533 cases, further case exclusions included 36 TBM cases with concurrent PTB and 17 PTB cases with lymph node enlargement / pleural pathology/ abnormal CT/MRI. Finally, 333 patients were classified as clinically confirmed TBM cases and 157 patients were classified as clinically confirmed PTB cases. Among these cases, those with negative cultures, HIV positive results and age ≤ 16 years of age at the time of diagnosis were excluded. Thus a total of 53 acid fast bacilli (AFB)/culture positive TBM cases and 72 AFB/culture positive PTB cases were selected, among which 10 archieved cultures for each group cases were processed for further analysis in each group. The study flow design is mentioned in **Figure 1**.

**Figure 1.**
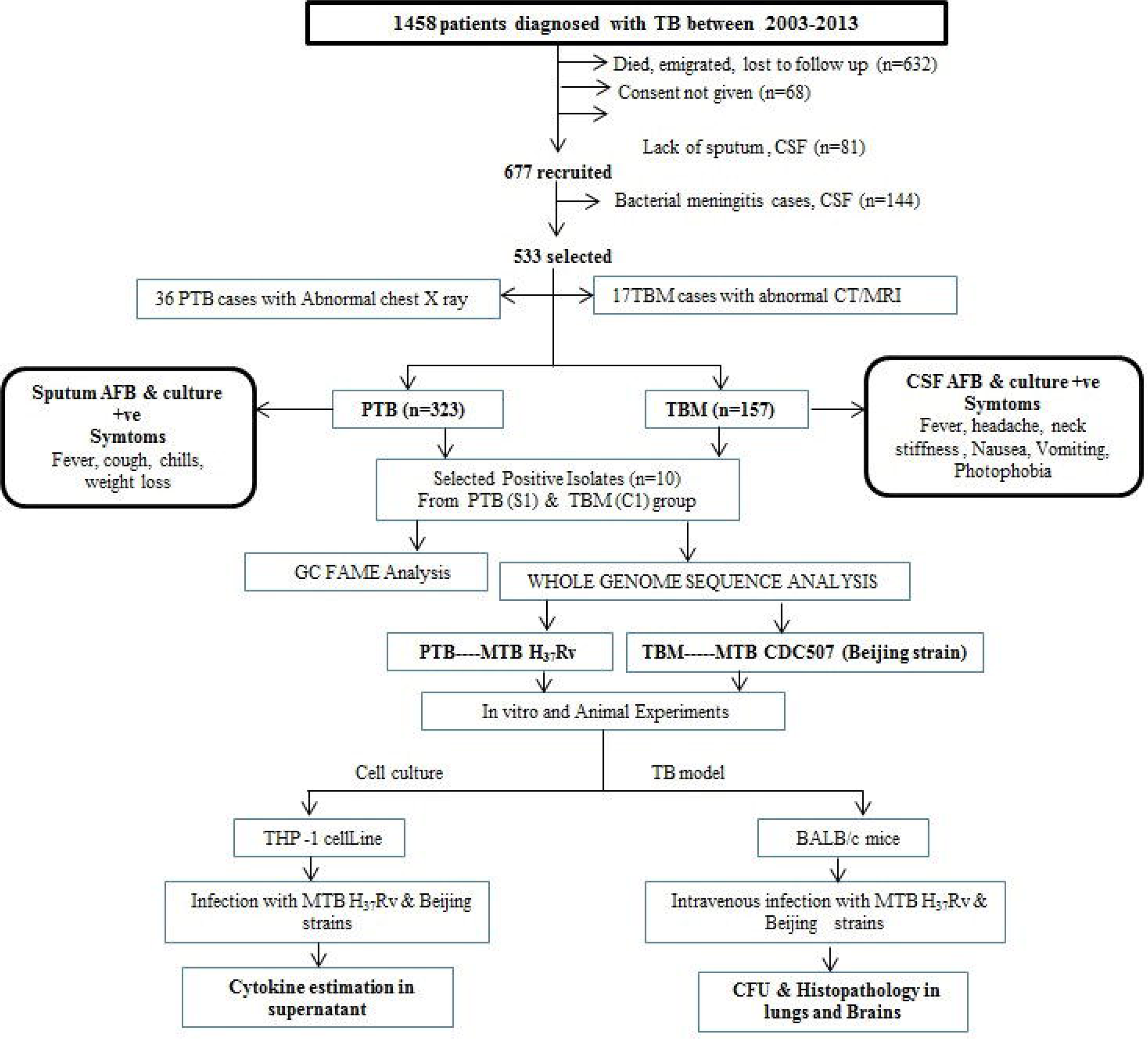
Study flow diagram for participant recruitment and experimental analysis

### 2.3 Sputum and CSF collection

2-3 ml of early morning sputum was collected in sterile sputum containers and immediately refrigerated at 4-8 °C until use. All samples were processed within 24-48 hrs after collection. CSF was collected by standard lumbar puncture techniques. 3-5mL of CSF was collected in sterile vaccutainer tubes from TBM patients admitted to the inpatient ward of CIIMS. Detection of AFB bacilli in both sputum and CSF was confirmed using Ziehl-Neelsen (ZN) staining and culturing in BacT/ALERT 3D automated culture system.

### 2.4 Culturing

2**-**3mL of sputum and CSF samples were cultured in Middle brook 7H9 liquid medium (Difco, USA) along with oleic acid, albumin, dextrose and catalase (OADC, Biomeriux, France) enrichment and antibiotic supplements in BacT/ALERT culture bottles (Biomeriux, France). These were incubated at 37°C in the BacT/ALERT 3D system (Biomerieux, France) for 28-35 days

### 2.5 Gas chromatographic analyses of Fatty Acid Methyl Esters (GC FAME)

For GC-FAME, 2-3 mL broth from positive cultures of both strains (n=10, each strain) were subcultured into a 100 mL flask containing Dubos liquid media (Hi Media Lab, India), supplemented OADC enrichment (BD Biosciences, USA) and 0.5 % glycerol and incubated for 10-15 days at 37°C until growth reached O.D_600_ 1.0. The fatty acids were extracted and methylated to form fatty acid methyl esters (FAME) as per procedure described elsewhere [11]

### 2.6 Whole genome shotgun sequencing

Whole genome shotgun sequencing of selected MTB isolates (S3 and C3 group) was performed on an IlluminaMiSeq platform with more than 100X sequence coverage. For microbial identification, DNA was amplified with a 27F primers set and sequenced using ABIBigDye terminator chemistry. The sequences were BLAST against nr data bases. The pair-end sequencing library was prepared using *illuminaTruSeq DNA Library Preparation Kit*. Library preparation was started with gDNA fragmentation of 1µg DNA, followed by pair-end adapter ligation. The ligated product was purified using 1X Ampure Beads. The purified ligated product was subjected to size-selection at ∼400-650 bp. The size-selected product was PCR amplified as described in the kit protocol. The amplified libraries were analyzed in Bioanalyzer 2100 (Agilent Technologies) using High Sensitivity (HS) DNA chip as per manufacturer’s instructions. After obtaining the concentration of the library with Qubit Fluorometer and its mean peak size of Bioanalyzer profile, 10 pM of the library was loaded onto MiSeq for cluster generation and sequencing. Sequencing was done using the 2 x 250 bp (500 cycles) chemistry to generate the raw data.

### 2.7 Genome Analysis

After sequencing, the genome was submitted to the Rapid Annotation using Subsystem Technology (RAST) server for functional annotation and genome comparison [12]. The genome data was submitted to National Center for Biotechnology Information (NCBI) with C3 and S3 reference number CP023170 and CP023169, respectively. Orthologous Average Nucleotide Identity Tool [13] was used to measure the overall similarity between four of the genomes (MTB strain H37Rv, CCDC5079, C1 and S1 strain). Mauve software was used for genome alignment and finding nucleotide level differences between the two genomes [14]. The genomes were further compared with a reference genome i.e. H_37_Rv (accession no. NC_018143) strain for S3 and CCDC5079 (accession no. NC_021251) for C3 using Briggs software, where H_37_Rv & CCDC5079 were taken as the reference genome [15]. Pfam was employed for domain analysis of the exclusive proteins found in each genome, for which the protein sequence of each protein in fasta format was submitted to the pfam server [16]. PE-PGRS and PE-PPE sequences were manually extracted, filtered, sorted and aligned using Geneious software [17]. Geneious software was also used for finding the least conserved region in the 4 MTB strains displaying deletion and SNP variation. Determination of 3D structure of the protein was carried by Swiss-Model server (homology based modelling) and visualized by UCSF Chimera using pdb file generated by the Swiss-model server [18-19]

### 2.8 Monocyte preparation and MTB infection

The human monocytic leukemia cell line, THP-1 was obtained from National Center for Cell Sciences (NCCS) Pune, India and cultured in RPMI 1640 (Gibco) supplemented with 2mM L-glutamine and 10 % fetal bovine serum (Sigma). Monocytes were exposed to respective MTB strains at a multiplicity of infection (MOI) in 1:1 ratio. After 2 hours of infection, the cells were gently washed three times with RPMI 1640 and treated with gentamycin (50ug/mL) to kill extracellular bacteria followed by centrifugation at 600 × g for 10 min to selectively spin down cells. Cells were resuspended in complete medium and cultured for 48 hours. Cell supernatant was collected at 48 hours and centrifuged for 10 min at 1000 g to get rid of cells and debris. The supernatant was stored at −20°C until assayed for cytokines. Each experiment was repeated in triplicate.

### 2.9 Cytokines estimation

Cytokines (IL-6, TNF-α, and VEGF) were assayed in cell supernatants by quantitative ELISA assay (Bender Med Systems, USA) as per manufacturer’s instructions.

### 2.10 Experimental mice

Inbred female BALB/c mice aged 6–10 weeks were procured from the National Institute of Immunology, New Delhi, India. Animals were housed in a Biosafety Level 3 facility at the JALMA, Agra, India. Before experiments all animal were acclimatized to housing condition for 14 days and fed with water and food under asceptic conditions.

### 2.11 Development of MTB infection and estimation of mycobacterial burden

For development of TB infection, two groups of experimental mice (n=12) were infected intravenously through the tail vein with 2×10^7^ CFU with both MTB strains. A control group of mice (n=10) without infection was separately maintained. Mice from each group (n=4) were sacrificed at 30 and 50 days after infection for CFU analysis and histopathology as per protocol described elsewhere [20].

### 2.12 Statistical Analysis

The frequencies (percentage) of demographic, clinical and risk factors were measured on a nominal scale. Comparison between disease groups were performed using the Chi square test in MedCalc statistical software (version 10.1.2.0) and a difference with p<0.05 was considered to be significant. All experimental results are expressed as mean ± standard deviation (SD) and presented on linear scale except for CFU, where logarithmic scale has been used. All statistical analysis were performed using Prism 6 version 6.01, GraphPad software, (San Diego, CA). Difference in the cytokines level, CFU analysis between experimental and control group was determined by t test. Test for proportion was used for comparison of total mortality in the experimental and control group. P value < 0.05 was considered statistically significant

## 3. Results

### 3.1 Baseline characteristics

Out of 1458 eligible participants, 323 cases satisfying the primary inclusion criteria were considered for the study. The baseline characteristics of the study population is presented in **Table 1**. The median age of PTB patients (58.8 years) was lower than that of TBM patients (63.9 years) (p < 0.001). We also examined the age distributions stratified by TB type. Our results confirmed the tendency for TBM and PTB to occur at age ≥ 40 years. The overall male to female ratio of TB cases was 1.7 (302/ 178). For TBM and PTB patients, the male to female ratio was 2 (216/107) and 1.2 (86/71) respectively. The difference was however not statistically significant. History of TB in the past was more common among PTB patients (5.13% vs. 1.2%). The proportion of patients who had a history of contact with a known case of TB (26.8% vs. 6.8%) was significantly higher among PTB patients. A significantly higher proportion of PTB cases had a cough and fever as compared to TBM cases (P<0.0001). On the other hand, a higher proportion of TBM cases had headache, neck stiffness, seizures, limb weakness and reduced level of consciousness (P<0.01). The mean duration of illness was also significantly higher in TBM cases as compared to PTB cases (P<0.0001). As compared to PTB patients a significantly higher proportion of TBM patients had concomitant diabetes mellitus (3.8% vs. 9.6%) (p < 0.05). A higher percentage of PTB cases had malignancies as compared to PTB cases, which did not reach statistical significance. The proportion of patients who were hypertensive (17.9% vs. 7.6%) was significantly higher among the TBM cases as compared to PTB cases (p < 0.05 ). The proportion of patients who were either currently smoking or ex-smokers was significantly higher among PTB patients (54.1%) compared to TBM patients (23.8%) (P < 0.0001). Similarly, a higher proportion (i.e. 34.4%) of PTB patients were either currently consuming alcohol or former alcohol drinkers (Table 1), and this difference was statistically significant (p < 0.05)

**Table 1.** Baseline & clinical characteristics of the 323 EPTB and 157 PTB participants under study: Percentages in different categories are indicated in parentheses.

| Sr. No. | Characteristics | TBM (n=323)<br>n (%) | PTB (n=157)<br>n (%) | P value |
| --- | --- | --- | --- | --- |
| 1 | Age, years, mean ± SD | 63.9 ± 17.3 | 58.8 ± 18.8 | **P < 0.0001** |
| 2 | Age groups, years |  |  |  |
|  | ≤ 18 | 6 (1.8) | 2 (1.3) | P = 0.9827 |
|  | 18-40 | 106 (32.8) | 73 (46.5) | **P = 0.0049** |
|  | ≥ 40 | 211 (65.4) | 82 (52.2) | **P = 0.0078** |
| 3 | Gender |  |  |  |
|  | Male | 216 (66.9) | 86 (54.8) | **P = 0.0133** |
|  | Female | 107 (33.1) | 71 (45.2) | **P = 0.0133** |
| 4 | History of TB | 4 (1.2) | 8 (5.1) | **P = 0.0234** |
| 5 | Family h/o TB | 22 (6.8) | 42 (26.8) | **P < 0.0001** |
| 6 | Cough | 0 (0) | 133 (84.7) | **P < 0.0001** |
| 7 | Fever | 166 (51.4) | 102 (65) | **P = 0.0066** |
| 8 | Headache | 232 (71.8) | 28 (17.8) | **P < 0.0001** |
| 9 | Neck stiffness | 222 (68.7) | 0 (0) | **P < 0.0001** |
| 10 | Seizures | 28 (8.7) | 2 (1.3) | **P = 0.0033** |
| 11 | Limb weakness | 74 (22.9) | 7 (4.5) | **P < 0.0001** |
| 12 | Reduced level of consciousness | 45 (13.9) | 3 (1.9) | **P = 0.0001** |
| 13 | Duration of illness (mean of days) | 172 | 161 | **P < 0.0001** |
| 14 | Clinical Outcome |  |  |  |
|  | Improved | 248 (76.8) | 144 (91.7) | **P = 0.0001** |
|  | Not improved | 62 (19.2) | 13 (8.3) | **P = 0.0032** |
|  | Expired | 13 (4) | 0 (0) | **P = 0.0251** |
| 15 | Diabetes mellitus | 31 (9.6) | 6 (3.8) | **P = 0.0400** |
| 16 | Malignancy | 4 (1.2) | 4 (2.5) | P = 0.5016 |
| 17 | Hypertension | 58 (17.9) | 12 (7.6) | **P = 0.0042** |
| 18 | Smoking | 77 (23.8) | 85 (54.1) | **P < 0.0001** |
| 19 | Alcoholism | 67/323 (20.7) | 54/157 (34.4) | **P < 0.0001** |

### 3.2 MTB isolates C1 and S1 exhibited differential growth pattern and fatty acid composition

Both C3 and S3 showed a difference in growth patterns. C3 isolates were associated with comparably higher growth times (43 days) as compared to S3 (29 days) which demonstrated rapid growth in BacT/ALERT 3D system (**Figure 2a**). The positive cultures from system (n=10, each) were then collected and used for GC-FAME analysis to study the unique fatty acid profiles of each strain. FAME profiles generated through GC-MS, subsequently compared using a microbial identification system are represented in **Figure 2 b**. Both MTB strains were associated with distinct FAME profiles. S3 showed significantly (P < 0.0001) higher levels of 10Me-18:0 TBSA FAME, with mean retention time of 15.955±5.27, compared to C3 (8.83±2.97). All C3 strain were associated with higher expression profiles of FAME 18:1w9c (17.555±7.35) compared to S3, although the variations were not significant statistically (**Figure 2 c**).

**Figure 2:**
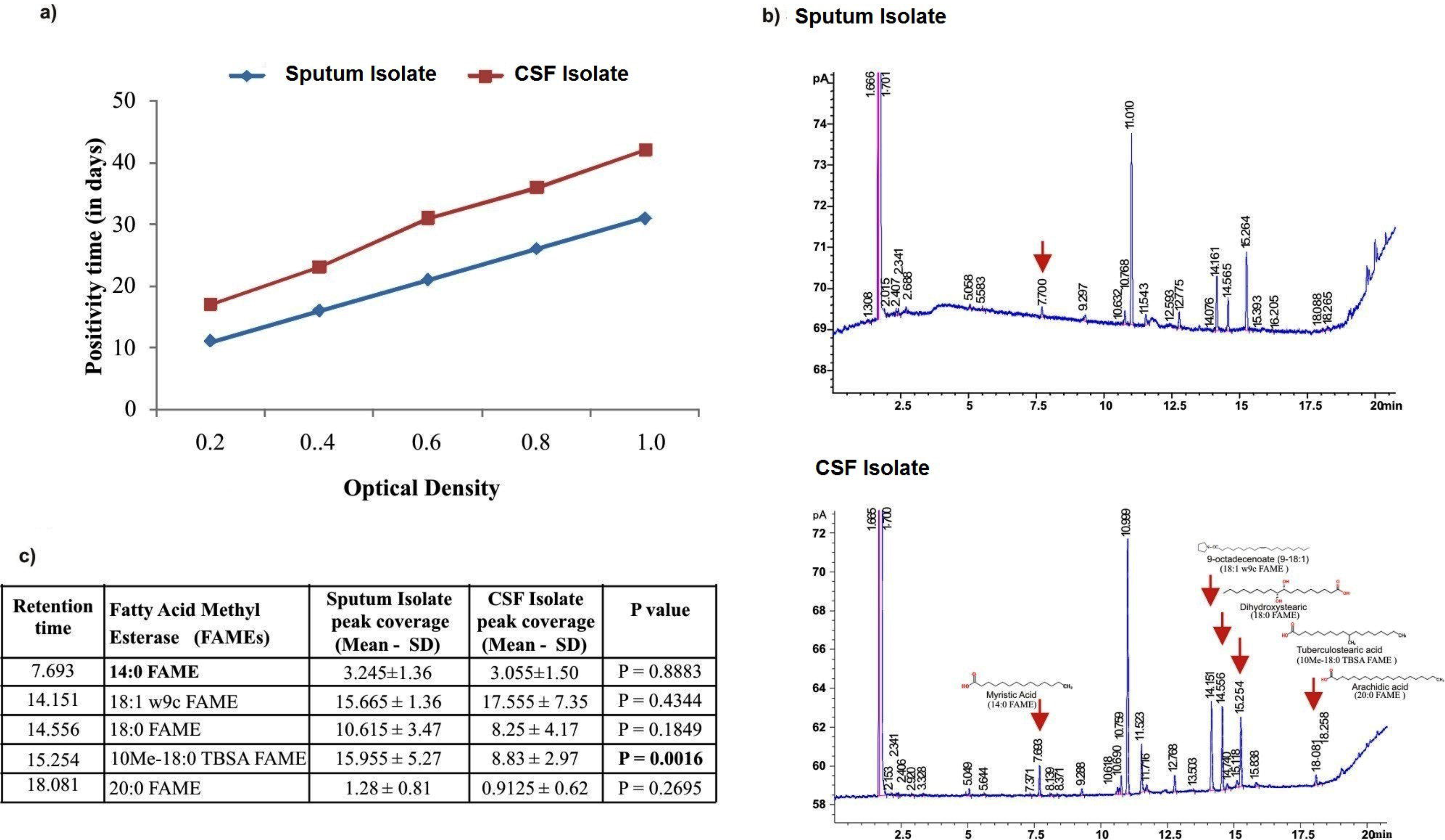
**a)** Growth pattern of two MTB isolates form CSF (C3) and Sputum (S3) samples cultured in BacT/Alert 3D system b) GC-FAME profiles of two MTB isolates along with fatty acid and alcohol designation. Arrows indicate differences in FAME profile of CSF isolate from sputum isolate. c) Characteristics of alcohol, fatty acid after GC FAME analysis associated with of MTB sputum and CSF isolates

### 3.3 Whole genome sequencing showed distinct genomic variation in derived from TBM and PTB cases

Total high quality data of 462 Mb and 1.08 Gb were generated for MTB S3 and C3 isolates using Illumina Miseq technology respectively. The mapping was performed on both isolates against MTB genomes using bwa 0.7.5a. This resulted in 1005066 mapped reads for C3 and 2518638 mapped reads for S3 with more than 99% of genome coverage on respective mycobacterium genomes (**Table 2**). The total gene sequences were extracted from consensus using the .gff file (from NCBI database) with the help of in-house perl scripts. This was followed by synteny analysis between C3 and S3. The C3 genome was identified as having a total of 4,406,296 bp with 4,204 genes and the S3 strain with 4,411,638 bp and 4,111 genes (**Table 2**). The mapping and alignment of the genome with representative genomes shows that C3 is 99.93% similar to MTB CCDC5079 (East—Asian/ Beijing family) and S1 is 99.96% similar to MTB H_37_Rv (Euro-American) (**Figure 3a**). The Blast analysis carried out by Brigg clearly indicate the high level of similarity existing between our and representative genomes (**Figure 3b**). The genes annotated by RAST subsystem shows differences in number of annotation (supplementary table 1) of which 24 genes can be seen in C3 and S3 out of which 9 genes were exclusively present in C3 strain and 17 genes exclusively present in S3 strain (**Table 3**). Comparative analysis in Mauve assist in highlighting the region where major differences in the genome exist (**Figure 4)**. Phage PhiRv1 genes (phage major capsid protein, phiRv1 prohead protease, probable phiRv1 phage protein, phage associated DNA helicase, putative phiRv1 phage protein, probable phiRv1 integrase and partial REP13E12 repeat protein) were found to be present in S3 but absent in C3 strain (**Figure 4a**). Five CRISPR genes (Cas2, Cas1, Csm6, Csm5 and Csm4) were also located in S3 which were absent from C3 strain (**Figure 4a**). A large deletion in C3 genome was also found at two of the locus where PE-PGRS virulence associated protein was present in S3 (**Figure 4b**).

**Table 2:** HQ data, Reference genome and Mapping statistics of CSF (C3) and Sputum isolate (S3) after Whole genome sequencing

| **Isolate** | **CSF isolate** | **Sputum Isolate** |
| --- | --- | --- |
| *Total number of bases (Mb)* | 462 Mb | 1.08 Gb |
| *Reference Genome* | CCDC5079 (no. ofbp=4414325) | H37Rv (no. of bp=4411708) |
| *Genome coverage (bp) without N's* | 4406296 | 4411638 |
| *% of Genome coverage* | 99.81 | 99.99 |
| *Total no. of reads* | 1013013 | 2536093 |
| *No. of mapped reads* | 1005066 | 2518638 |
| *No of unmapped reads* | 7947 | 17455 |
| *GFF file used* | CCDC5079. gff | H37Rv. Gff |
| *No. of genes (Chr.)* | 4204 | 4111 |

**Table 3:**
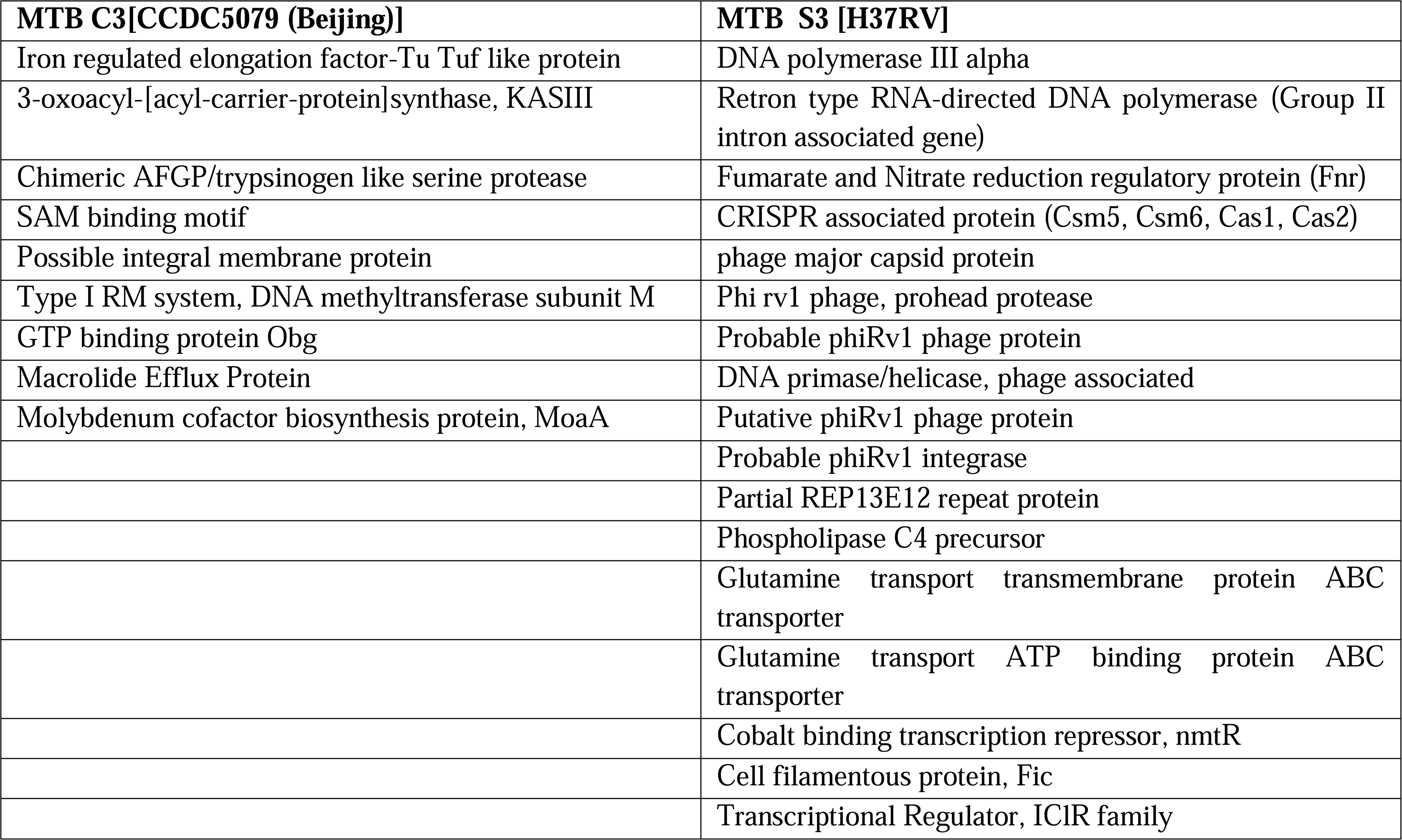
Genes exclusively present in respective genomes of the MTB C3 and S3 strain as identified after whole genome analysis

**Figure 3.**
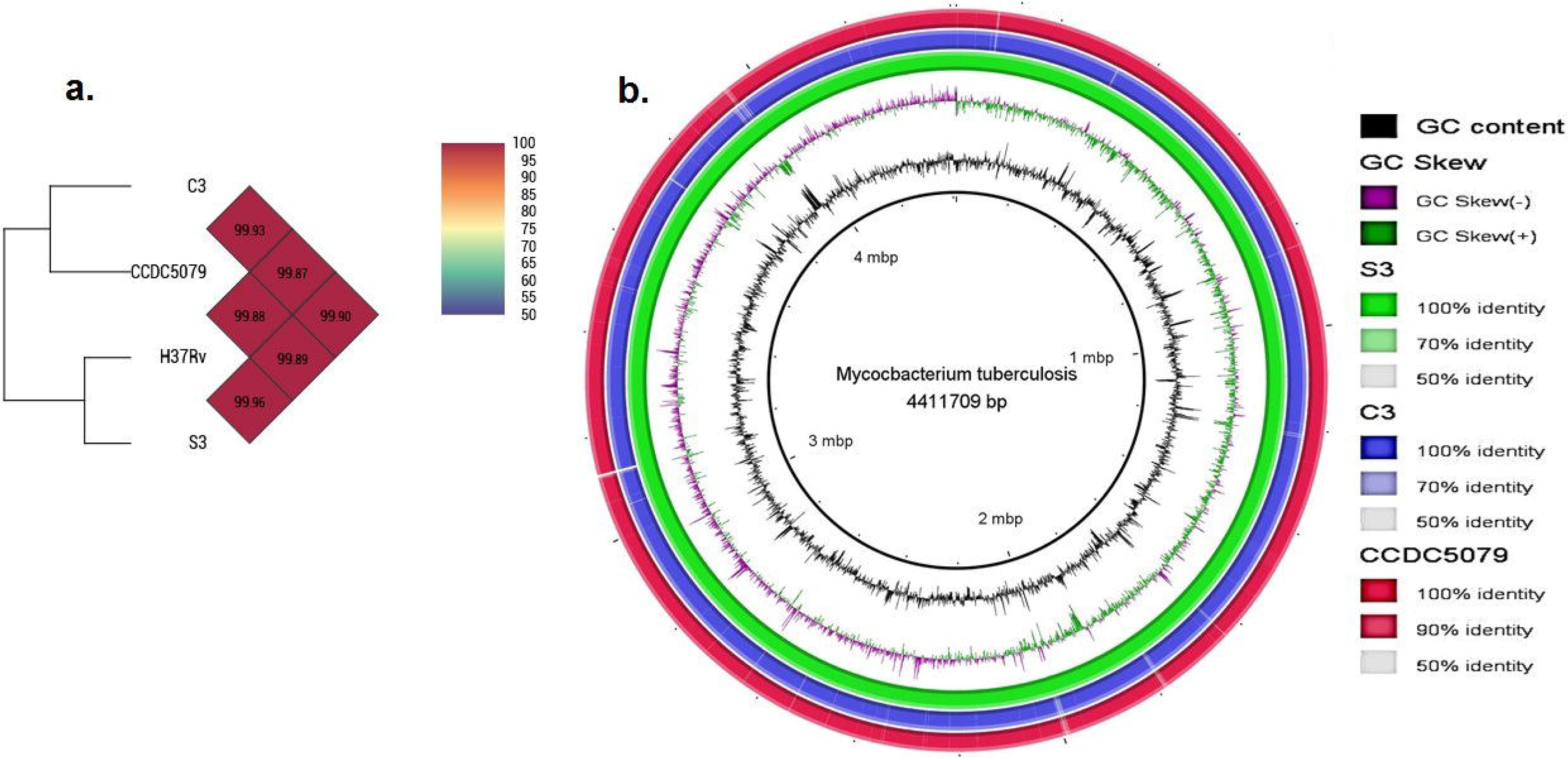
**a)** shows heatmap generated with OrthoANi values calculated from the OAT software. It clearly shows the genome closeness between C3 and CCDC5079 strain of MTB and S3 and H37Rv strain of MTB. The tree also shows that the two strain under study belongs to two different lineages of MTB. b) BLAST analysis of 4 *Mycobacterium tuberculosis* strains through BRIGG. Strain H37Rv was taken as reference genome. The green ring represent H37Rv vs S3, blue represent H37Rv vs C3 and red ring represent H37Rv vs CCDC5079

**Figure 4.**
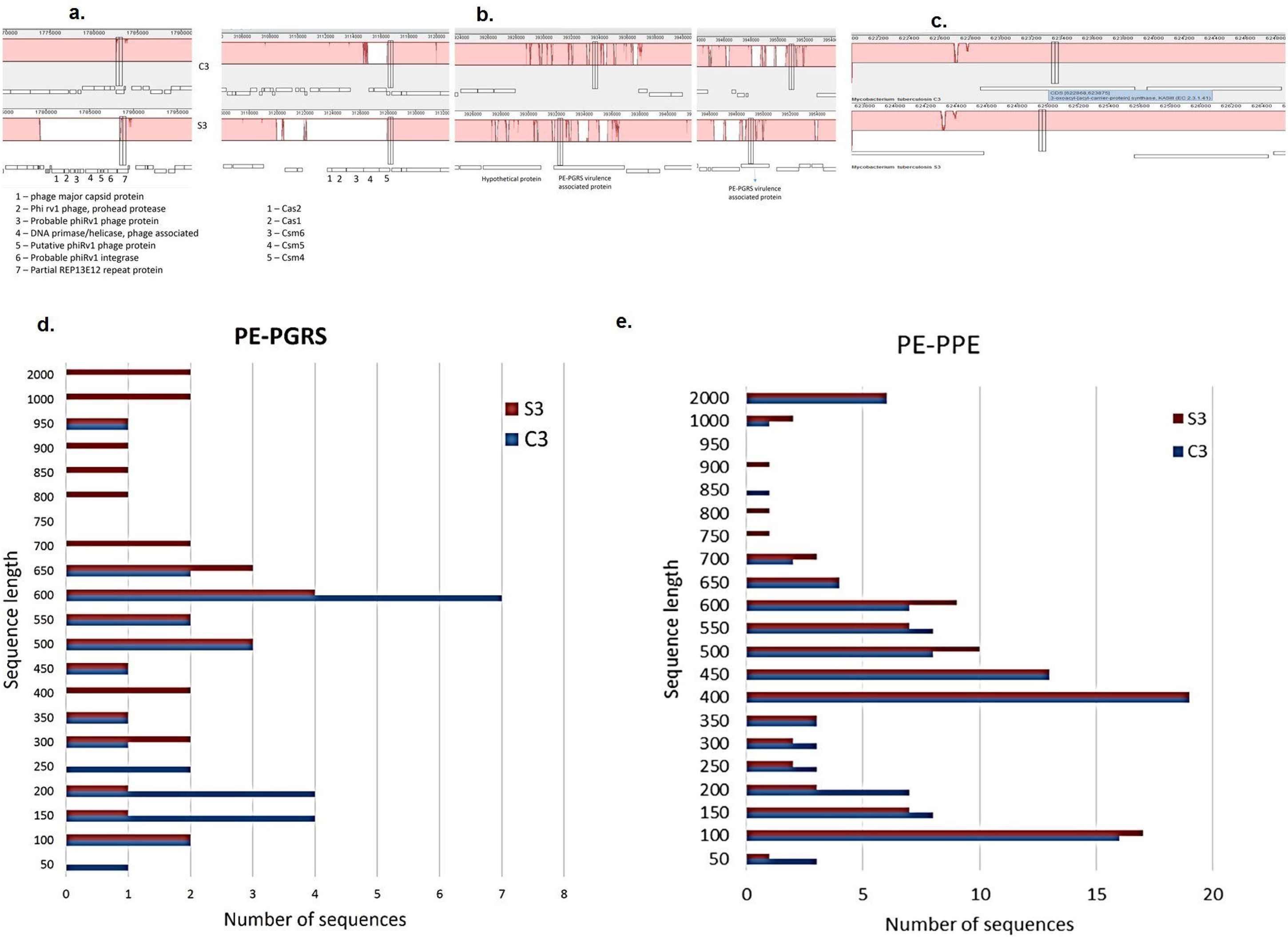
shows the comparative analysis between strain C3 and S3 at gene level generated through progressive alignment using Mauve software. a) indictes the genomic region where phage and CRISPR gene are present in S3 but not in C3. b.) The large deleted region in C3 is shown at the locus where PE-PGRS genes are present in S3. c) shows region where KASIII enzyme in present in C3 but absent in S3. d) variation of PE-PGRS and e) PE-PPE element existing in the two genome of M. tuberculosis strain C3 and S3. Geneious software was used for extraction, sortment and alignment of these elements. Finally the plot was generated using MS-Excel. More variation exist in case of PE-PGRs element than PE-PPE elements

### 3.4 Strain variations occur in membrane forming and membrane associated proteins

Genetic differences between the two strains were revealed when it was discovered that 3-oxoacyl-[acyl-carrier-protein (ACP)] synthase III was located in C3 but absent in S3 genome (**Figure 4 c**). Analysis in pfam confirmed that the two domains present in the protein belongs to ACP synthase III family. A back search in the genome through RAST confirmed that no other copy of this gene exists in the genome of either strains. Swiss-model server predicted the 3-dimensional structure of the MtfabH (3-oxoacyl-[acyl-carrier-protein (ACP)] synthase III) protein which in comparison with other MtfabH protein, shows 100% similarity (Supplementary Figure S1). When lipoproteins present in both the genomes were compared with the help of Pfam server, we discovered that lipoprotein LppE was truncated at the N-terminal in C3 and both the protein of LppA (Lipoprotein A) were also truncated in C3 at N-terminal (Supplementary Figure Figure S2 a). One of the desaturase related genes was found to be deactivated in the C3 genome by the insertion of the mobile element (Supplementary Figure S2 b).

### 3.5 PE-PGRS and PE-PPE are hotspot sites for mutations

The number and length of the PE-PGRS and PE-PPE proteins vary with the species and the strain. The plot shows that a large a amount of variation exists between both genomes in the case of the PE-PGRS element, with much less variation seen for PE-PPE (**Figure 4d & e**). S3 consisted of a PE-PGRS element of longer length than C3. Comparison of all the four MTB genome strains through geneious software showed that the site of PE-PGRS and PE-PPE element is a hotspot site of mutation with a large amount of deletions, single nucleotide variation (SNP) and insertions present in this region (Supplementary Figure S3).

### 3.6 Infection in THP-1 cell lines by both C3 and S3 showed a differential cytokine pattern

To study differential inflammatory response, cytokine expression profile induced by both C3 and S3 strains was studied using the THP-1 cell line model. **Figure 5 abc** shows mean cytokine levels in cell supernants of THP-1 cell lines after infection with both MTB strains. The C3 strain induced significantly higher levels (P>0.05) of inflammatory cytokines including TNF-α and VEGF compared to the S3 strain. Both strains were, however associated with similar expression levels of IL-6

**Figure 5:**
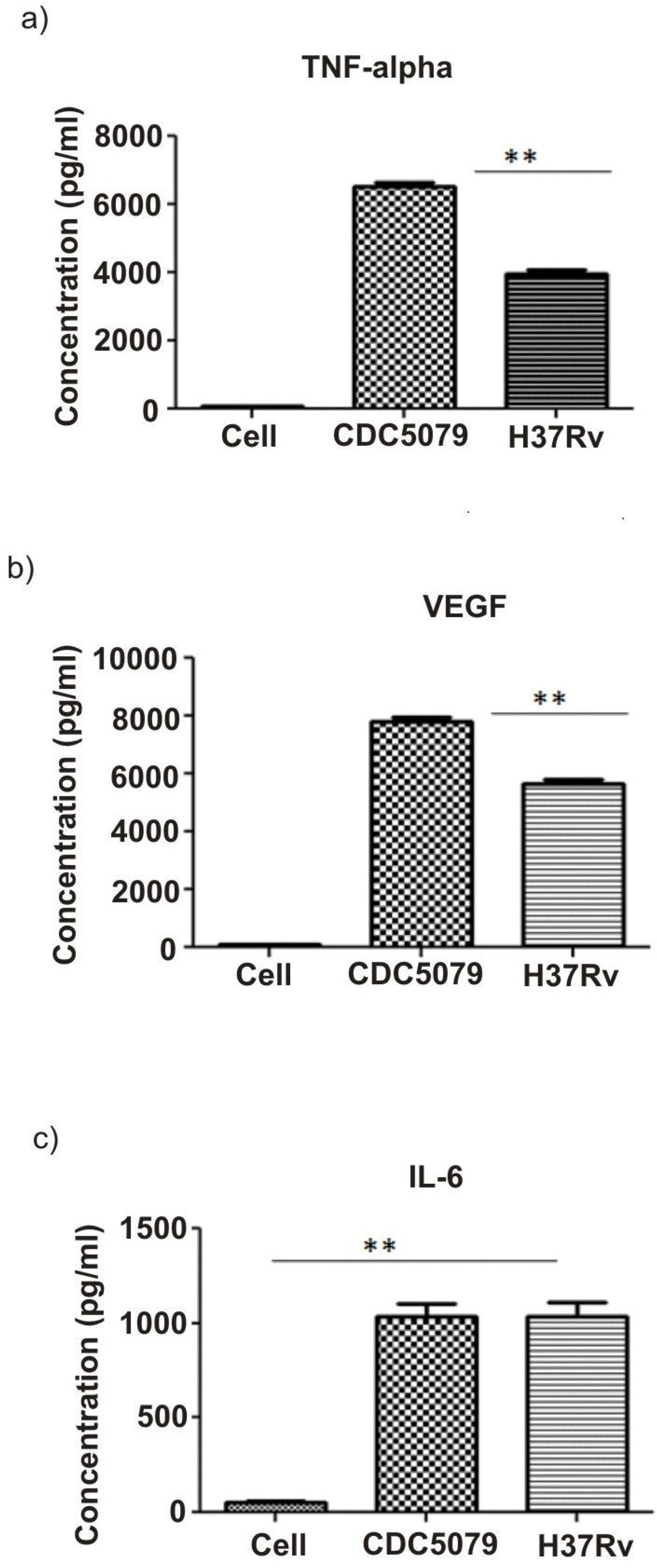
Cytokine profile a) TNF, b) VEGF and c) IL-6 in THP cell supernatants induced by C3 and S3 Strain. Monocytes were exposed with MTB at a multiplicity of infection (MOI) of one bacterium per cell ie. in1:1 ratio. Cell supernatant was collected at 48 hours and assayed for cytokines. Each experiment were repeated in triplicates. * represent statistically significant (P>0.05) and ** highly significant values (P>0.001). d & e)

### 3.7 Infection by the C3 strain in BALB/c mice was associated with more pronounce extra pulmonarydissemination and TBM development compared to S3 strain

To study pathogenesis and strain specific dissemination of both MTB strains, groups of mice were sacrificed at 30 and 50 days after infection and organs were isolated to examine lung and brain pathology. After isolation, organs were first visually examined for observation of amount of edema/swelling in the brain and granulomatous lesions in the lungs and brain (**figure 6 abc**). Mice infected with the CCDC5079 (C3) strain showed prominent swelling/edema of the brain in the left cerebral hemisphere at 30 days, which was further increased with progression in infection compared to control mice. Visual examination of brain of mice infected with the H_37_R_v_ strain however revealed no apparent abnormality with its morphology similar to the control mouse group. Examination of the lungs of mice infected with both MTB strains showed chronic pathology with multi-lobed lungs observed at 30 days. Lungs of mice infected with H_37_R_v_ (S3) strain showed progressive lung pathology with visibly evident granulomatous lungs at 50 days (**6a**). On the contrary, lungs of CCDC5079 infected mice showed gross reduction in pathology compared to H_37_R_v_ at 50 days (**6b**). No abnormality was observed in lungs and brains of control mice (a). Histopathological examination of the brain sections of both strain groups showed swelling of neurons along with lymphocytic infiltration which was progressively more prominent at 50 days in the CCDC5079 strain infected mice. Edema of brain parenchyma was also evident in brains of infected mice at both given time points after infection compared to H_37_R_v_ strain and control group (**6b&c**) which showed normal brain pathology with no evident infiltration and edema development at both 30 and 50 days. Examination of the lung sections revealed chronic pathology with high lymphocytic infiltration and an alveolar space mostly filled with inflammatory cells and edematous fluid in lung parenchyma of the pulmonary strain at 30 and 50 days post infection. Mice infected with CCDC5079 strain showed similar lung pathology at 30 days, but were associated with gross reduction in lung pathology with low infiltration and a largely preserved alveolar space with fewer cells filled with edematous fluid with progress in infection towards 50 days (**6b&c**).

**Figure 6.**
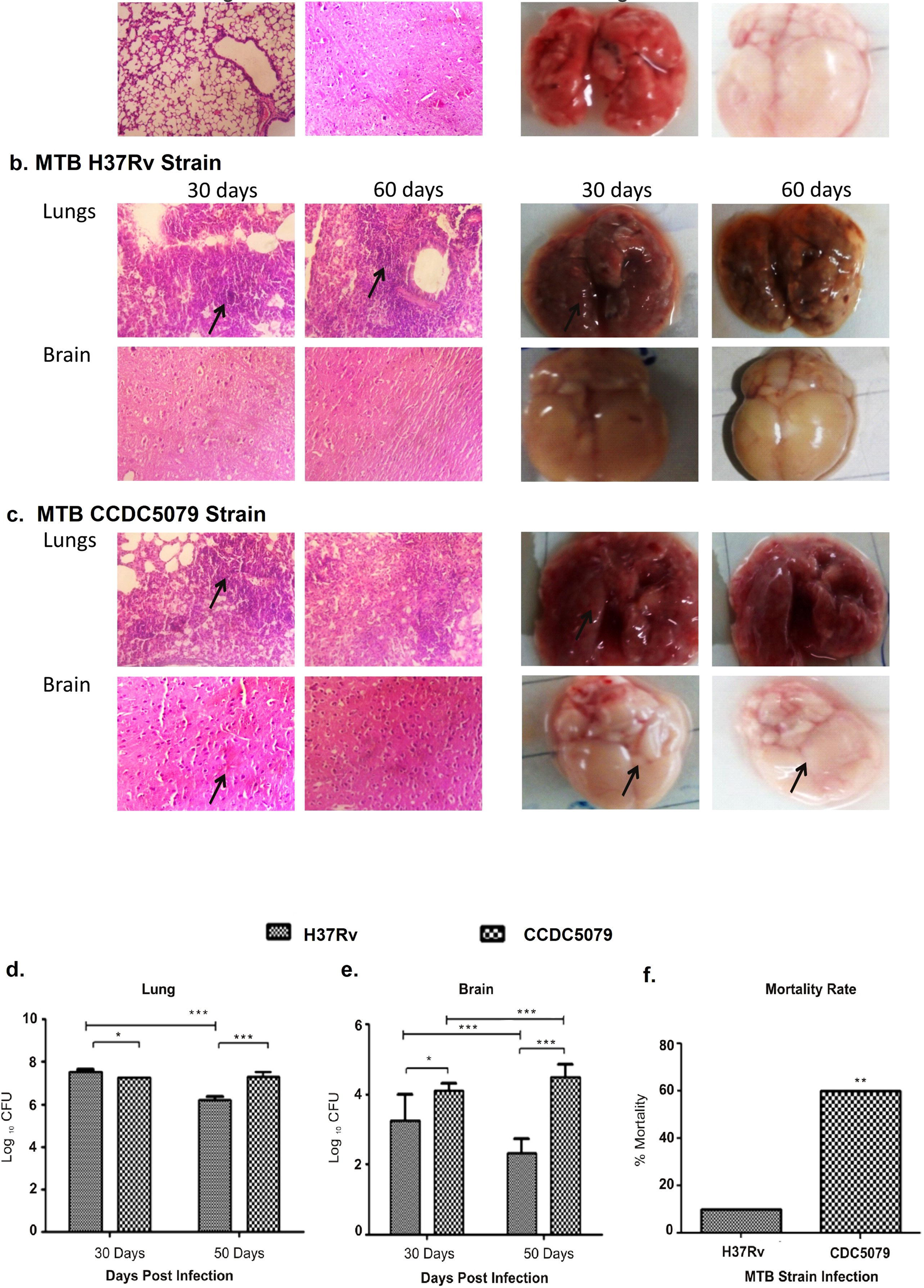
Representative photographs & histological examination lungs and Brain tissue of BALB/c mice 30 days and 50 days of a) control & mice post infection with b) MTB H_37_R_v_ (S3) and c) MTB CCDC5079 (C3) strain. Mice (n=4) were sacrificed 30 days and 50 days after intravenous infection with MTB strain. Lungs and brains of both mice group were examined visually as well by histological sectioning for gross pathological changes compared to control. Mice infected with S_3_ strain were associated with chronic lung pathology with high lymphocytic infiltration at both time points after infection. on contrary mice with infection of C3 strain showed chronic brain pathology with development of edema and swelling of neurons and lymphocytic infiltration. Both Strains of mice were associated with different organ pathology. Arrows indicate gross lung and brain pathology mice with MTB infection. Bacillary load in d) lungs and e) brains of mice infected with and C3 strain. 30 and 50 days post infection, mice (n=4) were sacrificed and organs were isolated, homogenized and serially diluted. 10 fold dilution of lung and brain homogenates were then were plated onto Middlebrook 7H11 medium with 0.05% Tween-80 and glycerol.. Bacillary burden in both organ are represented as log_10_ CFU. (**) represent significant (p<0.05) and (***) represents highly significant (p<0.001) values. f) Indicates % mortality rates associated with S3 and C3 strain post infection in mice

CFU analysis of both organs revealed significantly (P<0.05) higher bacterial load in the lungs (7.65±0.2) of H_37_R_v_ strain infected mice at 30 days post infection, compared to CCDC5079 strain (7.2±0.2) **(Figure 6d**). Analysis of CFU load in the brain showed significantly higher (P<0.001) mycobacterial load associated with mice infected with CCDC5079 compared to H_37_R_v_ strain at both given time points after infection (**Figure 6e**). Analysis of mortality rates associated with both strains revealed, significantly high mortality (60%) rate in CCDC5079 strain compared to H_37_R_v_ (20%) post infection (**Figure 6f)**

## 4. Discussion

The aim of the study was to investigate whether variation in MTB genotype or phenotype is responsible for differential site specific colonization in host leading to different disease outcome upon infection. To ascertain this, differences in genetic diversity in the mycobacterial strains and whole cell fatty acid composition were characterized using whole genome sequencing and FAME analysis, respectively. The MTB strains were further assessed using in vitro and murine MTB infection model studies to understand their infection dynamics and virulence patterns.

In recent years, various molecular phenotypes of MTB have been described and characterized through next genearation sequencing tools [21-22]. Reports from these studies have shown association of the genetic variations with mycobacterial pathogenesis and drug resistance [21]. As a consequence, these different genotypes have been found to induce variable host immune responses, which in turn may play a key role in manifestation of clinical characteristics.

Whole genome shotgun sequencing of MTB isolates in the present study demonstrated that both S3 and C3 isolates belong to different lineages/genotypes. Our data shows that the C3 strain which belongs to lineage 2 (East asian), has a propensity to exhibit a more highly virulent phenotype than Lineage 4 (Euro-American) of S3, favoring selection of highly virulent bacteria resulting in CNS dissemination [23]. Our results are in agreement with earlier studies that have found only the Beijing, Indo-Oceanic and Euro-American lineages from TBM patients [24]. Contrastingly, some authors have reported that clustered MTB isolates, albeit observed at low frequency, are found in TBM patients [25].

Although reports suggest that genetic diversity in MTB might contribute to disease presentation in extrapulmonary sites. [26], the results from our genomic analysis would appear to show a lack of extensive variation between the the two MTB isolates. Genes such as *ptpA, Rv1988* and *Rv1099c, PkND* reported in earlier studies to evoke differential immunological responses in the host [6,27-29] were present in both strains. This was further supported through Mauve analysis which did not reflect SNP variation in any of the important genes. Our results suggest that there is no direct relationship between host-pathogen interaction and specialized localization of two of the MTB strains. However, alteration in the membrane profile of the bacterial cells, including components of the cell membrane, cell wall, lipoproteins and receptor proteins might contribute to variation seen in how the strains interact with the host immune system, leading to differential localization of the two strains.

The presence of a full set of phage *phiRv1* genes and *CRSISPR* genes in S3 strain depicts a major difference between the two strains. However, studies have reported that the presence of phages in the MTB aids only in the evolution and adaptation and have no direct effect in the modulation of pathogenesis behavior of the organism [30]. The presence of *3-oxoacyl-[acyl-carrier-protein (ACP)] synthase III gene* in the C3 demarcates a major line between C3 and S3. This gene is responsible for production of beta-ketoacyl-ACP which on further processing in bacteria, plays a significant part in phospholipid biosynthesis and cell wall synthesis [30]. Fatty acid biosynthesis in bacteria consists of two systems; FAS-I (de novo synthesis) and the FAS-II system which is an elongation step. The mycobacterial KAS-III enzyme belongs to the FAS-II system and assists in fatty acid elongation by catalyzing the reaction between C_12_-CoA and Malonyl-ACP molecule [31]. Scarsdale et al [32] have reported this KASIII enzyme (mtFabH) in MTB and also pointed out its role in initiating meromycolic acid biosynthesis. On mtFabH protein structural analysis using Pfam and 3D structure, we have shown that mtFabH in C3 is 100% similar with that reported by Scarsdale et al. Thus, with the presence of mtFabH, the mycolic acid composition and cellular membrane profile of the C3 will be significantly variable in comparison with S3 where this enzyme was absent. This can be further supported with FAME anlaysis which shows the presence of different lengths of fatty acid methyl esterases in C3 which were absent in S3 isolate. Various reports have been published showing different genotypes of MTB are associated with difference in FAMEs [33-35]. Similarly, the differences seen in lipoprotein profiles may be variable between the two strains due to presence of altered lipoprotein LppE and LppA in C3. This can also have a major impact in altering lipoproteins, which are a crucial target for immune response and pathogenesis [36]. Additionally we found one of the Desaturase related proteins to be deactivated by mobile element in C3 and not in S3suggesting that altered membrane profile changes may lead to differential localization of the two MTB strains in the human host. Alongside these proteins, there was significant variation reported in PE-PGRS and PE-PPE elements of both strains. PE and PE-PGRS, of which both are localized to the cell membrane, are associated with host-pathogen interactions which play a significant role in virulence and modulation of immune response [37-38]. These elements were also reported as having adhesion related properties imparting its position in adhesion with the host cell surface [39]. Analysis of these elements on both C3 and S3 showcase large disparities in terms of number, length and nucleotide sequence. While the S3 features the long sequences of PE-PGRS in its genome, C3 is rich in a number of shorter PE-PGRS element. We also discovered that there are 3 PE-PGRS element that are exclusively present in S3 and absent from C3 and H37Rv genome., Our results reveal on comparative analysis of the strains that PE-PGRS and PE-PPE elements were the hot-spot sites of mutations. Therefore, changes in this element can widely alter the response mechanism of the host immune system towards mycobacterium strains leading to differential localization of this strain in human i.e. C3 in the brain and S3 in the lung. Adding to this the presence of plcD gene in S3 strains (absent in C3) emphasizes its localization in lung region. The plcD gene found in S3 is 99% similar with CDC1551 and this gene has been reported to have a role in pathogenesis of TB[40]

Post ascertaining and identification of two distinct clinical strains of MTB, infection dynamics of both C3 and S3 were studied using in vitro and animal model of TBM infection. In vitro studies using MTB infection with both C3 and S3 strain showed a diverse cytokine profile with significantly higher TNF & VEGF levels associated with C3 strains, compared to S3 strains. Further studies in animals with high dose intravenous infection with two MTB strains showed diverse infection dynamics with high extrapulmonary dissemination exhibited by C3 strain compared to S3. Both strains showed diverse organ pathology with higher brain pathology and CFU load in the brain associated with C3 strain compared to S3 strain which induced more pulmonary than meningeal infection. MTB, C3 was found to be more virulent due to CNS pathogenesis with higher mortality rates compared to S3. Earlier works by various investigators using vitro and animal studies have exhibited different behavior of the disease that was connected to the genotype of infecting bacteria. Chacón-Salinas R, et al investigated differential patterns of cytokine expression in macrophages infected with distinct MTB genotypes such as Canetti, Beijing, and H37Rv and found that each genotype has ability to induce different levels of cytokine expression [41]. Similar works by Rivera-Ordaz A et al in peripheral blood mononuclear cells (PBMC) of healthy BCG vaccinated individuals showed that MTB, Beijing strains showed differential cytokine production compared to H37RV with high TNF levels [42]. Other studies have shown that certain strains can adhere, invade and transverse endothelial cells by virtue of virulence factors and inducing cytokines like VEGF. VEGF has been significantly associated with TBM pathogenesis by participating in the breakdown of BBB permeability. [43].

In the past several years there have been numerous animal models that have investigated the virulence and pathogenesis post infection with different MTB genotypes [5,44-45]. In our earlier studies, we have specifically shown that high intravenous infection with C3 strain leads to development of CNS disease in mice, with high bacterial burden in brains peaking at 50 days post infection [18] In the present study, to ascertain, similar infection dynamics and to compare tissue specific colonization, we have used a similar infection model from our earlier studies using two different MTB strains. Our present results indicate tissue specific colonization which aligns with other reports which have shown that extrapulmonary dissemination in MTB infection is associated with strain specific virulence [46-49].

Above studies are associated with some limitation, which include limited MTB isolates from TBM and PTB patients for Whole genome and fatty acid analysis. Similarly, through in vitro studies, we were able to study limited cytokine profile, compared to multiple cytokine which could have given a better picture of microbial modulation of immune response. In the study, we have given limited focus on host factors that may also play an important role in diverse MTB pathogenesis.

To conclude, our study shows that dissemination and development of TBM and PTB disease may be attributed to MTB strain-specific differences. The study also acknowledges that variation in fewer enzymes can lead to differences in virulence property via differential membrane composition in the MTB. The above study will be helpful to rationalize our efforts in understanding strain specific virulence patterns of PTB and TBM disease for improved diagnostics and therapeutic interventions, which may be useful in reducing overall mortality associated with these diseases in developing regions.

## Data Availability

Available on request

## 5. Acknowledgement

The authors would like to thank Pathology department of the Nagpur Veterinary College for histopathological examination of brain and lungs section. Authors also acknowledge Colorado State University, USA, for providing MTB antigens and antibodies under the TB Research Materials and Vaccine Testing Contract (NO1-AI-75320). Authors acknowledges support of Mrs Anju Mudliar for FAME analysis, Ms Seema Shekhawat for technical support in cell line study and Clininicans Dr Dinesh Kabra and Dr Neeraj Baheti for providing patient details and clinical management in recruited cases. Hitesh Tikariha acknowledges the fellowship received from Uinoversity Grants Commission. All authors acknowledge CIIMS, Nagpur for funding this study as part of in house research grant.

## 7. Author contributions

Conceptualization (AAH, UDG, NCC, HFD, RSK), Formal analysis (AAH, UDG, ARN, SDB, HJP,RSK) Data curation (AAH, HT, SDB, HJP, RSK), Investigation (AAH, HT, ARN, UDG, SDB, NCC, HJP, RSK), Methodology (AAH,HT,ARN,UDG,SDB,NCC,HJP, RSK), Project administration (AAH, RSK), Supervision (AAH, HFD, RSK), Validation (AAH, HJP, RSK, UDG), Funding acquisition (HFD, LRS, RSK), Resources (LRS, RSK), Writing – original draft (AAH, HT,) Writing – review & editing (AAH, ARN, NCC, TMM, HFD, LRS, HJP, RSK)

## References

1. Kashyap RS, Husain AA. Over-the-counter drug distribution and tuberculosis control in India. Lancet Infect Dis 2016; 16:1208–09.

2. Murthy JM. Tuberculous meningitis: the challenges. Neurol India 2010; 58:716–22.

3. Be NA, Kim KS, Bishai WR, Jain SK. Pathogenesis of central nervous system tuberculosis. Curr Mol Med 2009; 9:94–9.

4. Rock RB, Olin M, Baker CA, Molitor TW, Peterson PK. Central nervous system tuberculosis: pathogenesis and clinical aspects. Clin Microbiol Rev 2008; 21:243–61

5. Isabel BE, Rogelio HP. Pathogenesis and immune response in tuberculous meningitis. Malays J Med Sci 2014; 21:4–10

6. Jain SK, Paul-Satyaseela M, Lamichhane G, Kim KS, Bishai WR. Mycobacterium tuberculosis invasion and traversal across an in vitro human blood-brain barrier as a pathogenic mechanism for central nervous system tuberculosis. J Infect Dis 2006; 193:1287–95

7. Tsenova L, Ellison E, Harbacheuski R, Moreira AL, Kurepina N, Reed MB, Mathema B, Barry CE 3rd, Kaplan G. Virulence of selected Mycobacterium tuberculosis Clinical isolates in the rabbit model of meningitis is dependent on phenolic glycolipid produced by the bacilli. J Infect Dis 2005; 192:98–106.

8. Brown T, Nikolayevskyy V, Velji P, Drobniewski F. Associations between Mycobacterium tuberculosis strains and phenotypes. Emerg Infect Dis 2010; 16: 272–80.

9. Garcia de Viedma D, Marin M, Ruiz Serrano MJ, Alcala L, Bouza E. Polyclonal and compartmentalized infection by Mycobacterium tuberculosis in patients with both respiratory and extrarespiratory involvement. J Infect Dis 2003; 187:695–9.

10. WHO. Standards for TB care in Inida, 2014 [internet]. Available at http://www.tbcindia.nic.in/showfile.php?lid=3061 [Accessed 2019 December]

11. Ozbek A, Aktas O. Identification of three strains of Mycobacterium species isolated from clinical samples using fatty acid methyl ester profiling. J Int Med Res 2003; 31:133–40.

12. Overbeek R, Olson R, Pusch GD, Olsen GJ, Davis JJ, Disz T, Edwards RA, Gerdes S, Parrello B, Shukla M, et al. The SEED and the Rapid Annotation of microbial genomes using Subsystems Technology (RAST). Nucleic acids research 2013; 42:D206–14.

13. Lee I, Kim YO, Park SC, Chun J. OrthoANI: an improved algorithm and software for calculating average nucleotide identity. International journal of systematic and evolutionary microbiology. 2016;66:1100–3.

14. Darling AC, Mau B, Blattner FR, Perna NT. Mauve: multiple alignment of conserved genomic sequence with rearrangements. Genome research. 2004; 14:1394–403.

15. Alikhan NF, Petty NK, Zakour NL, Beatson SA. BLAST Ring Image Generator (BRIG): simple prokaryote genome comparisons. BMC genomics 2011;12:402.

16. Finn RD, Coggill P, Eberhardt RY, Eddy SR, Mistry J, Mitchell AL, Potter SC, Punta M, Qureshi M, Sangrador-Vegas A et al. The Pfam protein families database: towards a more sustainable future. Nucleic acids research 2016;44:D279–85.

17. Kearse M, Moir R, Wilson A, Stones-Havas S, Cheung M, Sturrock S, Buxton S, Cooper A, Markowitz S, Duran C, et al. Geneious Basic: an integrated and extendable desktop software platform for the organization and analysis of sequence data. Bioinformatics. 2012;28:1647–9.

18. Biasini M, Bienert S, Waterhouse A, Arnold K, Studer G, Schmidt T, Kiefer F, Cassarino TG, Bertoni M, Bordoli L, et al. SWISS-MODEL: modelling protein tertiary and quaternary structure using evolutionary information. Nucleic acids research. 2014; 42:W252–8.

19. Pettersen EF, Goddard TD, Huang CC, Couch GS, Greenblatt DM, Meng EC, Ferrin TE. UCSF Chimera—a visualization system for exploratory research and analysis. Journal of computational chemistry. 2004; 25:1605–12.

20. Husain AA, Gupta UD, Gupta P, Nayak AR, Chandak NH, Daginawla HF, Singh L, Kashyap RS. Modelling of cerebral tuberculosis in BALB/c mice using clinical strain from patients with CNS tuberculosis infection. Indian J Med Res 2017; 145:833–839

21. Advani J, Verma R, Chatterjee O, Pachouri PK, Upadhyay P, Singh R, Yadav J, Naaz F, Ravikumar R, Buggi S et al. Whole Genome Sequencing of Mycobacterium tuberculosis Clinical Isolates From India Reveals Genetic Heterogeneity and Region-Specific Variations That Might Affect Drug Susceptibility. Front Microbiol 2019; 10:309.

22. Brown AC, Bryant JM, Einer-Jensen K, Holdstock J, Houniet DT, Chan JZ, Depledge DP, Nikolayevskyy V, Broda A, Stone MJ et al. Rapid Whole-Genome Sequencing of Mycobacterium tuberculosis Isolates Directly from Clinical Samples. J Clin Microbiol. 2015; 53:2230–2237.

23. Ribeiro SC, Gomes LL, Amaral EP, Andrade MR, Almeida FM, Rezende AL, Lanes VR, Carvalho EC, Suffys PN, Mokrousov I et al. Mycobacterium tuberculosis strains of the modern sublineage of the Beijing family are more likely to display increased virulence than strains of the ancient sublineage. J Clin Microbiol 2014; 52:2615–24.

24. Yorsangsukkamol J, Chaiprasert A, Prammananan T, Palittapongarnpim P, Limsoontarakul S, Prayoonwiwat N. Molecular analysis of Mycobacterium tuberculosis from tuberculous meningitis patients in Thailand. Tuberculosis (Edinb) 2009; 89:304e9.

25. Faksri K, Drobniewski F, Nikolayevskyy V, Brown T, Prammananan T, Palittapongarnpim P, Prayoonwiwat N, Chaiprasert A. Epidemiological trends and clinical comparisons of Mycobacterium tuberculosis lineages in Thai TB meningitis. Tuberculosis (Edinb) 2011; 91:594–600.

26. Sharma K, Verma R, Advani J, Chatterjee O, Solanki HS, Sharma A, Varma S, Modi M, Ray P, Mukherjee KK, et al. Whole Genome Sequencing of Mycobacterium tuberculosis Isolates From Extrapulmonary Sites. OMICS 2017; 21:413–425.

27. Wang J, Ge P, Qiang L, Tian F, Zhao D, Chai Q, Zhu M, Zhou R, Meng G, Iwakura Y et al. The mycobacterial phosphatase PtpA regulates the expression of host genes and promotes cell proliferation. Nat Commun 2017; 8:244

28. Yaseen I, Kaur P, Nandicoori, VK, Khosla S. Mycobacteria modulate host epigenetic machinery by Rv1988 methylation of a non-tail arginine of histone H3. Nature communications 2015; 6: 8922.

29. Ganapathy U, Marrero J, Calhoun S, Eoh H, de Carvalho LPS, Rhee K, Ehrt S. Two enzymes with redundant fructose bisphosphatase activity sustain gluconeogenesis and virulence in Mycobacterium tuberculosis. Nat Commun 2015;6:7912.

30. Kremer L, Dover LG, Carrère S, Nampoothiri KM, Lesjean S, Brown AK, Brennan PJ, Minnikin DE, Locht C, Besra GS. Mycolic acid biosynthesis and enzyme characterization of the beta-ketoacyl-ACP synthase A-condensing enzyme from Mycobacterium tuberculosis. Biochem J 2002; 364:423–30.

31. Brown AK, Sridharan S, Kremer L, Lindenberg S, Dover LG, Sacchettini JC, Besra GS. Probing the mechanism of the Mycobacterium tuberculosis beta-ketoacyl-acyl carrier protein synthase III mtFabH: factors influencing catalysis and substrate specificity. J Biol Chem 2005;280:32539–47.

32. Scarsdale JN, Kazanina G, He X, Reynolds KA, Wright HT. Crystal structure of the Mycobacterium tuberculosis β-ketoacyl-acyl carrier protein synthase III. Journal of Biological Chemistry 2001;276: 20516–22.

33. Mosca A, Russo F, Miragliotta L, Iodice MA, Miragliotta G. Utility of gas chromatography for rapid identification of mycobacterial species frequently encountered in clinical laboratory. J Microbiol Methods 2007; 68:392–5

34. Zerbini E, Cardoso M, Sequeira M, Taher H, Santi N, Larpin D, Latini O, Tonarelli G. Characterization of fatty acids and mycolic acid degradation products in mycobacterial species of major incidence in Argentina. Rev Argent Microbiol 1997; 29:184–94

35. Müller K, Schmid EN, Kroppenstedt RM. Improved identification of mycobacteria by using the microbial identification system in combination with additional mtrimethylsulfonium hydroxide pyrolysis. J Clin Microbiol 1998; 36: 2477–80.

36. Wilkinson KA, Newton SM, Stewart GR, Martineau AR, Patel J, Sullivan SM, Herrmann JL, Neyrolles O, Young DB, Wilkinson RJ. Genetic determination of the effect of post-translational modification on the innate immune response to the 19 kDa lipoprotein of Mycobacterium tuberculosis. BMC Microbiol 2009; 9:93.

37. Sampson SL. Mycobacterial PE/PPE proteins at the host-pathogen interface. Clin Dev Immunol 2011; 2011:497203

38. Fishbein S, Wyk N, Warren RM, Sampson SL. Phylogeny to function: PE/PPE protein evolution and impact on Mycobacterium tuberculosis pathogenicity. Molecular microbiology 2015; 96: 901–16.

39. Ramsugit S, Pillay M. Identification of Mycobacterium tuberculosis adherence-mediating components: a review of key methods to confirm adhesion function. Iranian journal of basic medical sciences 2016; 19:579.

40. Yang Z, Yang D, Kong Y, Zhang L, Marrs CF, Foxman B, Bates JH, Wilson F, Cave MD. Clinical relevance of Mycobacterium tuberculosis plcD gene mutations. Am J Respir Crit Care Med 2005;171:1436–42.

41. Chacón-Salinas R, SerafÍn-López J, Ramos-Payán R, Méndez-Aragón P, Hernández-Pando R, Van Soolingen D, Flores-Romo L, Estrada-Parra S, Estrada-GarcÍa I. Differential pattern of cytokine expression by macrophages infected in vitro with different Mycobacterium tuberculosis genotypes. Clin Exp Immunol 2005;140:443–9

42. Rivera-Ordaz A, Gonzaga-Bernachi J, SerafÍn-López J, Hernández-Pando R, Van Soolingen D, Estrada-Parra S, Estrada-GarcÍa I, Chacón-Salinas R. Mycobacterium tuberculosis Beijing genotype induces differential cytokine production by peripheral blood mononuclear cells of healthy BCG vaccinated individuals. Immunol Invest 2012;41:144–56..

43. Misra UK, Kalita J, Singh AP, Prasad S. Vascular endothelial growth factor in tuberculous meningitis. Int J Neurosci 2013; 123:128–32.

44. Zucchi FC, Tsanaclis AM, Moura-Dias Q Jr, Silva CL, Pelegrini-da-Silva A, Neder L, Takayanagui OM. Modulation of angiogenic factor VEGF by DNA-hsp65 vaccination in a murine CNS tuberculosis model. Tuberculosis (Edinb) 2013;93: 373–80.

45. Hernández Pando R. Modeling of Cerebral Tuberculosis: Hope for Continuous Research in Solving the Enigma of the Bottom Billion’s Disease. Malays J Med Sci 2011; 18:12–5

46. Be NA, Lamichhane G, Grosset J, Tyagi S, Cheng QJ, Kim KS, Bishai WR, Jain SK. Murine model to study the invasion and survival of Mycobacterium tuberculosis in the central nervous system. J Infect Dis 2008;198:1520–8

47. Brown T, Nikolayevskyy V, Velji P, Drobniewski F. Associations between Mycobacterium tuberculosis strains and phenotypes. Emerg Infect Dis 2010; 16:272–80.

48. Garcia de Viedma D, Marin M, Ruiz Serrano MJ, Alcala L, Bouza E. Polyclonal and compartmentalized infection by Mycobacterium tuberculosis in patients with both respiratory and extra respiratory involvement. J Infect Dis 2003; 187:695–9.

49. Hernandez Pando R, Aguilar D, Cohen I, Guerrero M, Ribon W, Acosta P, Orozco H, Marquina B, Salinas C, Rembao D, Espitia C. Specific bacterial genotypes of Mycobacterium tuberculosis cause extensive dissemination and brain infection in an experimental model. Tuberculosis (Edinb) 2010;90: 268–77.

