## Supplementary table 1 for "Variation in Mycobacterium tuberculosis genotype and molecular phenotype influence clinical phenotype of Pulmonary tuberculosis and Tuberculous Meningitis infection in host"

**Supplementary Table 1**: Genes Annoted by RAST in C3 and S3 genomes

|  | C3 | S3 |
| --- | --- | --- |
| Amino Acids and Derivatives | 314 | 315 |
| Carbohydrates | 289 | 290 |
| Cell Division and Cell Cycle | 44 | 43 |
| Cell Wall and Capsule | 75 | 73 |
| Cofactors, Vitamins, Prosthetic Groups, Pigments | 304 | 304 |
| DNA Metabolism | 93 | 101 |
| Dormancy and Sporulation | 2 | 2 |
| Fatty Acids, Lipids, and Isoprenoids | 244 | 245 |
| Iron acquisition and metabolism | 2 | 2 |
| Membrane Transport | 55 | 54 |
| Metabolism of Aromatic Compounds | 13 | 13 |
| Miscellaneous | 31 | 31 |
| Motility and Chemotaxis | 2 | 2 |
| Nitrogen Metabolism | 24 | 25 |
| Nucleosides and Nucleotides | 83 | 83 |
| Phages, Prophages, Transposable elements, Plasmids | 2 | 5 |
| Phosphorus Metabolism | 44 | 44 |
| Photosynthesis | 0 | 0 |
| Potassium metabolism | 12 | 12 |
| Protein Metabolism | 238 | 239 |
| Regulation and Cell signaling | 102 | 100 |
| Respiration | 99 | 98 |
| RNA Metabolism | 69 | 70 |
| Secondary Metabolism | 1 | 1 |
| Stress Response | 84 | 85 |
| Sulfur Metabolism | 34 | 36 |
| Virulence, Disease and Defense | 117 | 117 |
