## Supplementary figure 1 for "Variation in Mycobacterium tuberculosis genotype and molecular phenotype influence clinical phenotype of Pulmonary tuberculosis and Tuberculous Meningitis infection in host"

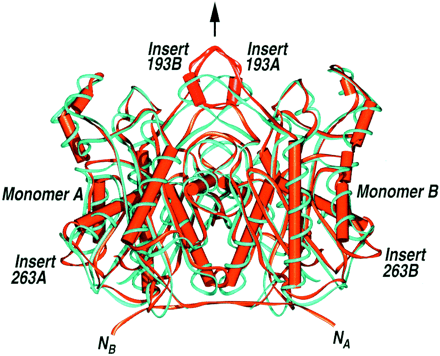

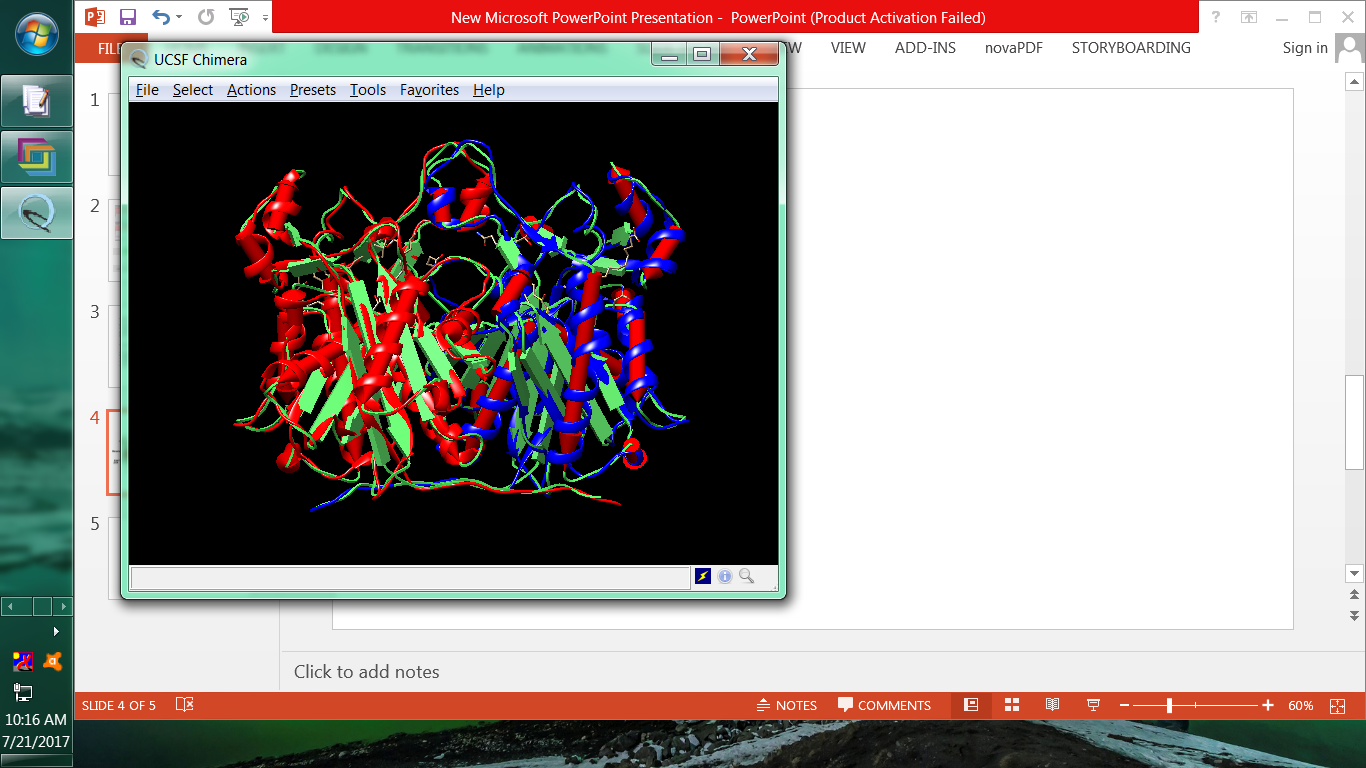

**Supplementary Figure S1**. The 3-dimensional structure of C3 MtfabH protein (left side) and Mtfabh obtained by Brown et al. (2005)(right side) shows 100% similarity. Swiss-Prot was used for determining 3D structure of mtFabh and UCSF Chimera software was used for visualization of generated protein 3D structure.
