## Supplementary figure 2 for "Variation in Mycobacterium tuberculosis genotype and molecular phenotype influence clinical phenotype of Pulmonary tuberculosis and Tuberculous Meningitis infection in host"

**Supplementary Figure S2**

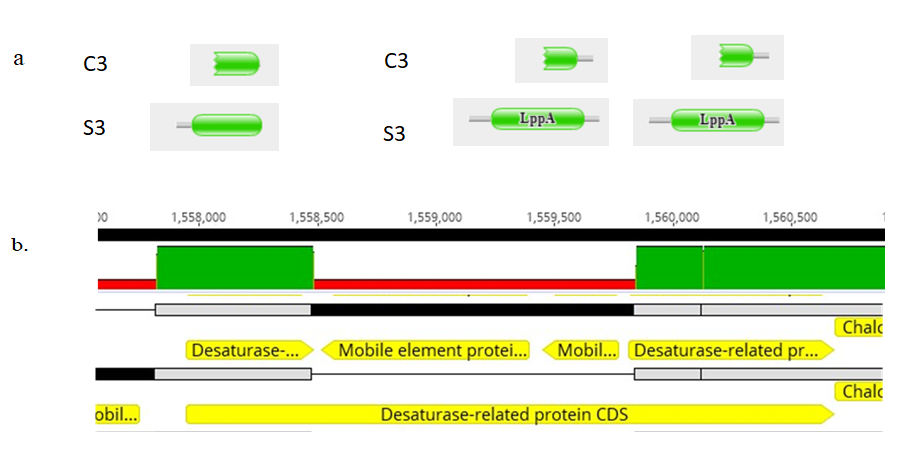

**Supplementary Figure S2. a.** Figure shows the domain level comparison of the lipoprotein present in M. tuberculosis C3 and S3. Pfam was used for domain determination. On the Left side is lipoprotein E (LppE) and on the right side is two of the lipoprotein A (LppA). **b.** The figure shows the region in genome of C3 where insertion of mobile element have inactivated one of the desaturase related protein whereas in S3 it is intact. The figure was generated through Geneious software by alignment of the two genome and zooming to show the gene level differences.
