## Supplementary figure 3 for "Variation in Mycobacterium tuberculosis genotype and molecular phenotype influence clinical phenotype of Pulmonary tuberculosis and Tuberculous Meningitis infection in host"

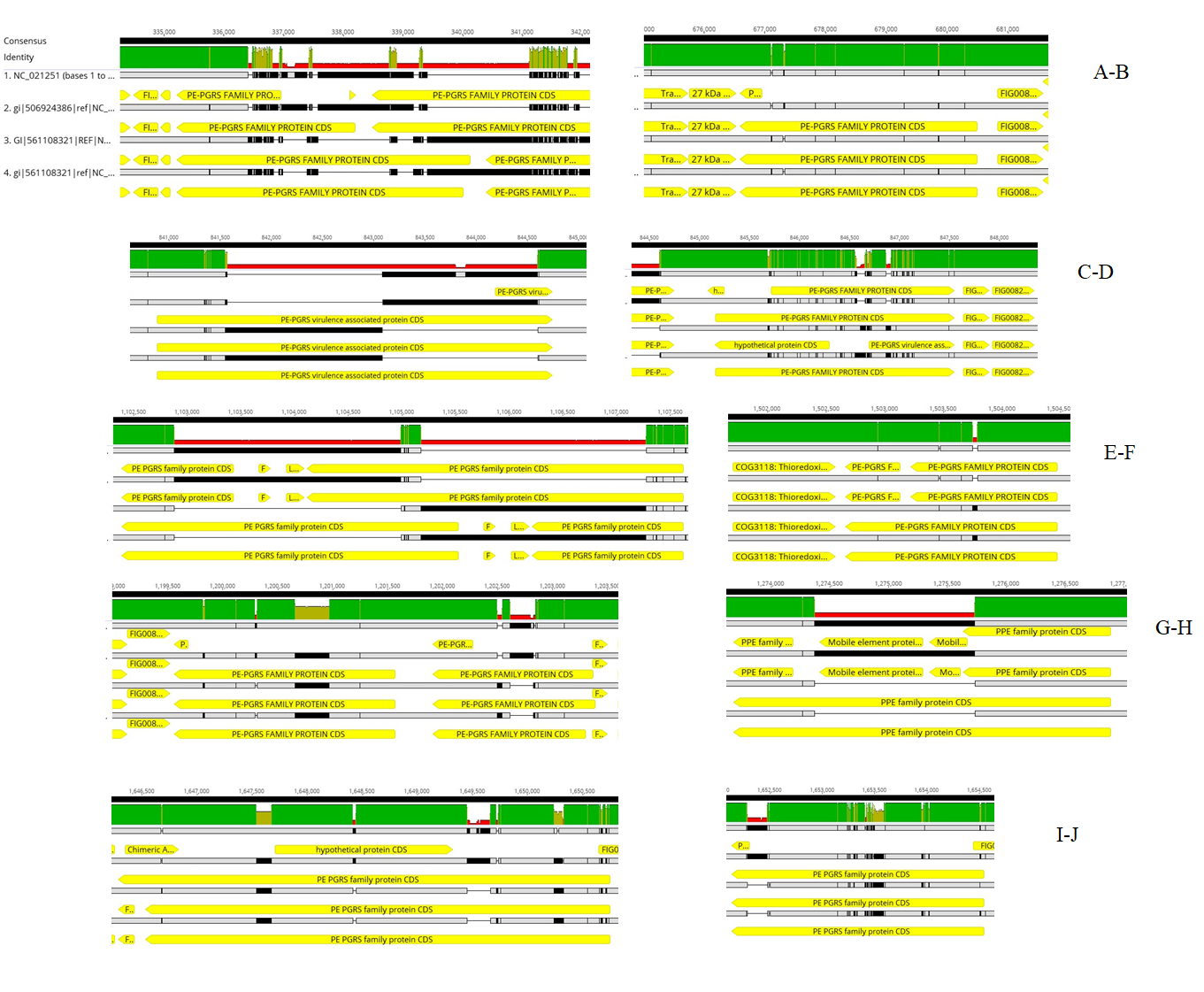

**Supplementary Figure S3**. Figure shows the PE-PGRS sites in all the 4 genome of M. tuberculosis strains linked with heavy mutagenesis, deletion and SNP variations. Figures are generated by whole genome alignment using Geneious software. From top to bottom, row 1: CCDC5079, row 2: C3, row 3: S3 and row 4: H37Rv.
